# The Mediterranean Diet is Associated with Higher Arterial Elasticity over Prefrontal Cortex in Older Adults

**DOI:** 10.64898/2026.04.20.26351341

**Authors:** Felicity Simpson, Jenna Johnson, Patrycja Kałamała, Monica Fabiani, Karen J. Murphy, Alexandra Wade, Ashlee Harvey, Nicholas Ware, Montana Hunter, Maddison L Mellow, Daniel Barker, Clare E Collins, Kathy Low, Gabriele Gratton, Hannah A.D. Keage, Ashleigh E. Smith, Frini Karayanidis

**Author notes:** **Corresponding authors:** Felicity Simpson, Frini Karayanidis.

## Abstract

**INTRODUCTION:** Healthful dietary patterns may attenuate dementia risk by preserving cerebrovascular health. Prior work has focused on systemic arterial stiffness, but cerebrovascular measures may be more sensitive to neuroprotective effects of diet. We examined associations between Mediterranean diet adherence, prefrontal cortex (PFC) arterial elasticity, and cognition in older adults.

**METHODS:** Participants were 198 older adults (58% female; mean age 65.6 years) from the Newcastle ACTIVate cohort. Mediterranean Diet (MedDiet) scores were derived from the Australian Eating Survey food frequency questionnaire. Pulse Relaxation Function (PReFx), an index of PFC arterial elasticity, was measured using pulse Diffuse Optical Tomography. Cognition was assessed with CANTAB and a cued task-switching paradigm.

**RESULTS:** Higher MedDiet was associated with higher PFC arterial elasticity. MedDiet was not associated with cognition, and PReFx did not mediate diet-cognition associations.

**DISCUSSION:** Greater Mediterranean diet alignment was cross-sectionally associated with PFC arterial elasticity, suggesting a pathway through which diet may influence brain health in ageing.

## 1. Introduction

As the global population ages, dementia prevalence is projected to reach 150 million cases worldwide by 2050 [1]. This rise in cases has prompted an emphasis on lifestyle-based prevention strategies, such as a balanced diet, to promote healthy ageing and delay the onset of dementia. Several studies have shown that consuming a healthful dietary pattern during mid- and late adulthood is associated with slowed cognitive decline and lower dementia risk [2]. However, the mechanisms by which healthful dietary patterns may preserve cognitive function in older adults are not fully elucidated. One plausible mechanism involves the vascular system, as healthy arteries support the maintenance of sufficient blood flow to the brain. Yet, to date, few studies have examined the association between dietary habits and measures of cerebrovascular health (see reviews: [3, 4]).

The Mediterranean diet (MedDiet) is characterised by high consumption of fruits, vegetables, whole grains, nuts, legumes, extra virgin olive oil (EVOO), and fish and low intake of discretionary foods. It is linked to reduced risk of chronic conditions, including cardiovascular disease and cognitive decline [5]. In a recent meta-analysis including 23 cohort studies, the MedDiet was associated with an 18% lower risk of cognitive impairment (13 studies), an 11% lower risk of dementia (10 studies) and a 30% lower risk of Alzheimer’s disease (9 studies)[6]. The MedDiet is also particularly notable for its association with cardiometabolic risk in both experimental and observational studies, with higher MedDiet associated with lower blood pressure, obesity, fasting glucose, inflammatory markers, and more optimal lipid profiles (see reviews: [7]).

A higher burden of these cardiovascular risk factors is associated with poorer cognitive performance and an increased risk of dementia [8]. Preclinical arterial changes, including reduced elasticity of arterial walls (arteriosclerosis) and the buildup of plaque that narrows the arterial shaft (atherosclerosis), cause vessels to become stiffer and less compliant, contributing to elevated blood pressure, which in turn further accelerates arterial stiffening [9]. As arterial stiffness persists, it initiates a cascade of damaging effects on the network of tiny vessels perfusing the brain, driving accelerated brain ageing [10]. Notably, studies using carotid-femoral pulse wave velocity (PWV), the gold standard measure of systemic arterial stiffness, have shown associations with cognitive decline (for review, see [11]).

Consuming a poor-quality diet may increase arterial stiffness, which, in turn, can elevate cerebral arterial pulsatility (the rhythmic variation in blood flow with each heartbeat) and impair cerebral perfusion, processes that have been implicated in cognitive decline [12]. Healthful dietary patterns have been consistently associated with lower arterial stiffness in middle-aged and older adults [13, 14, 15, 16]. Consistent with these findings, a meta-analysis of 38 trials found small to moderate reductions in arterial stiffness following dietary interventions [17].

To date, no study has directly examined the association between dietary patterns and cerebral arterial health. PWV measures stiffness in major bodily arteries but cannot resolve the smaller cerebral vessels responsible for brain perfusion, and intracranial Doppler is limited in older adults by calcification of the temporal acoustic window. The Pulse Relaxation Function (PReFx), derived from the arterial pulse wave using Diffuse Optical Tomography (pulse-DOT), indexes cerebral arterial elasticity by capturing how effectively cerebral arteries recoil after systolic expansion [18]. Lower PReFx has been associated with older age, lower cardiopulmonary fitness, greater volume of white matter hyperintensities, greater brain atrophy, and lower fluid intelligence [19, 20, 21, 22, 23].

This is the first study to investigate the relationship between the MedDiet and cerebral arterial elasticity. Specifically, we examine whether the MedDiet is associated with elasticity of cerebral arteries perfusing the prefrontal cortex, a region supporting executive functioning, one of the cognitive domains most sensitive to ageing [24]. We hypothesise that a higher MedDiet score will be associated with greater cerebral arterial elasticity (PReFx). We further hypothesise that the MedDiet will be positively associated with cognitive functioning, mediated by PReFx. Understanding these relationships may inform dietary strategies for preserving cerebrovascular health.

## 2. Methods

### 2.1 Participants

Cross-sectional data from the Newcastle arm of the ACTIVate research project [25] was used to investigate the association between diet, cerebral arterial elasticity, and cognition in older adults. The ACTIVate study was conducted in accordance with the Declaration of Helsinki, following approval obtained from the University of South Australia Research Ethics Committee (approval number: 202639) and co-registered with the University of Newcastle Human Research Ethics Committee. The study was registered with the Australian New Zealand Clinical Trials Registry (registration number: ACTRN12619001659190).

In total, 200 participants aged 60-70 years were recruited from the Newcastle site and screened over the phone (see Figure 1). Screening was conducted using the Telephone-Montreal Cognitive Assessment (T-MoCA) to ensure cognitive functioning was above the Mild Cognitive Impairment dementia threshold (>13/22 [26]). Additional exclusion criteria included drug or alcohol dependency, uncorrected vision problems, lack of fluency in English, stroke or transient ischemic attack, physical or intellectual disability, or cancer treatment in the last 5 years (see Smith et al. [25] for full study protocol).

**Figure 1.**
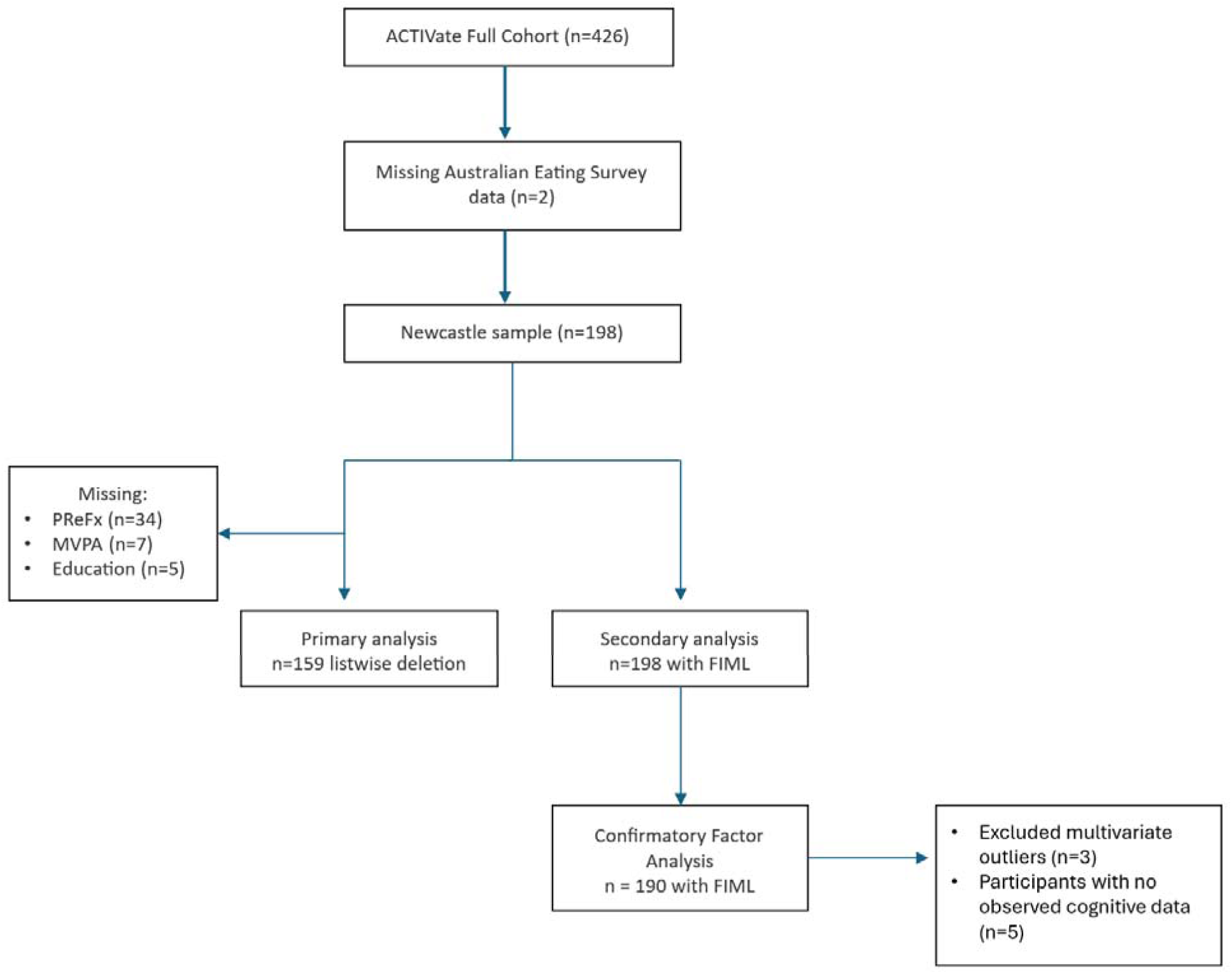
Flow Diagram for Final Participant Sample *Note*. The Newcastle analytic cohort comprised 198 participants. The primary regression sample was derived by listwise deletion, yielding n = 159 complete cases. For the CFA, cognitive indicators were screened for univariate outliers cell-wise (|z| > 3; 16 cells replaced with NA) and multivariate outliers via partial Mahalanobis distance (*p* = .003 cutoff); three participants were excluded, leaving n = 195. Five participants with no observed cognitive data were ignored by lavaan, yielding n = 190 for model estimation. Mediation models were estimated on the full cohort (n = 198) using FIML.

### 2.2 Materials

#### 2.2.1 Assessment of Diet

The Australian Eating Survey Food Frequency Questionnaire (AES-FFQ) is a self-report dietary assessment tool that asks individuals to report on consumption of 120 food items over the last six months [27]. Participants rate the frequency of consumption using a Likert scale ranging from ‘*Never’ to ‘4 or more times per day*’ for food items and to ‘*7 or more glasses per day’* for beverages, with each response level based on Australian food portion sizes.

The AES-FFQ has been evaluated for reliability and validity against a 3-day weighed food record (WFR). The intraclass correlation coefficient (ICC) against WFR for total energy intake was 0.60 (95% CI 0.43, 0.77), with a median ICC of 0.47 across all nutrients [26]. The AES-FFQ has also demonstrated validity in measuring fruit and vegetable intake compared with blood-based biomarkers including plasma and skin carotenoid concentrations [28, 29]. Nutrient intakes are derived using the AUSNUT 2011–2013 national nutrient database [30].

##### Mediterranean Diet (MedDiet) score

The MedDiet is typically characterised by the consumption of vegetables and fruit, EVOO, fish, grains and cereals, alongside a moderate consumption of alcohol and dairy foods, with minimal consumption of processed foods and red meat [5]. The MedDiet score was calculated using the Panagiotakos et al. [31] scoring system which includes 11 food categories (i.e., non-refined cereals, fruits, vegetables, legumes, potatoes, olive oil, fish, red meat, poultry, full-fat dairy products, and alcohol). Consumption frequencies for each category ranged from 0 (no consumption) to 5 (daily consumption), with red meat, poultry and full-fat dairy products scored inversely. Alcohol consumption below 300 mL/day was assigned the highest score of 5, while higher consumption (300–699 mL/day) received progressively lower scores (4–1 points); a score of 0 was assigned for either no consumption or consumption of 700 mL/day or more. The total MedDiet score ranged from 0 to 55, with higher scores indicating greater alignment to a MedDiet eating pattern.

#### 2.2.2 Assessment of Cerebral Arterial Health

The Pulse Relaxation Function (PReFx) was calculated to measure arterial elasticity using pulse-DOT data acquired over the frontal lobes. PReFx quantifies the shape of the cerebral arterial pulse waveform, based on changes in near-infrared light absorption related to haemoglobin oxygenation [18, 32]. The following sections describe the digitisation of source and detector positions, optical and ECG recording procedures, and MRI acquisition used to support co-registration and PReFx calculation.

##### Digitisation

A custom-made foam helmet was fitted to each participant’s head to hold the optical sources and detectors in place. The spatial locations of all source and detector positions were recorded using a Polhemus 3Space FASTRAK 3D digitisation system (Polhemus, Colchester, VT), with a stylus inserted into each designated point on the helmet. This was done so that source and detector positions could be realigned with each participant’s brain anatomy. Three anatomical reference landmarks (i.e., nasion, left and right preauricular points) were also digitised to optimise surface registration. Three head-mounted receivers allowed for minor head movement during digitisation. The stylus was calibrated using RealTerm software, and Locator software was used to map the scalp. Participants were instructed to avoid large head movements, and the helmet was not removed between digitisation and optical recording.

##### Optical recording

Optical data were recorded using a multi-channel frequency-domain oximeter (Imagent; ISS Inc., Champaign, IL, USA) with 16 time-multiplexed light emitting diodes and four detectors. Near-infrared light at 690 nm and 830 nm was delivered to the scalp via optical fibres (400 µm diameter), with laser diodes modulated at 110 MHz. Detector fibre bundles (3 mm diameter) collected the returning light and transmitted it to photomultiplier tubes (PMTs), which were modulated at 110.003125 MHz, generating a 3.125 kHz cross-correlation frequency. A Fast Fourier Transform (FFT) of the PMT output was used to extract direct current (DC), alternating current (AC) intensity, and relative phase. Optical parameters were sampled at 39.06 Hz (25.6 ms per sample).

Data were acquired from three consecutive montages covering the left, middle, and right frontal cortex as well as the anterior parietal and temporal lobes. For each montage, four 3-minute blocks were collected, comprising a resting block, two breath-holding task blocks, and a second resting block. The breath-holding data are not reported here.

##### Electrocardiogram (ECG) recording and analysis

The ECG was recorded time-locked with the optical recording, to ensure that the same pulse wave was measured at different points in the brain. The ECG was recorded using a PowerLab system (AD Instruments) with electrodes placed on the left inner elbow and both wrists. The signal was sampled at 1000 Hz and band-pass filtered between 1–50 Hz to minimise high-frequency noise and baseline drift. The ECG recording was used to time-lock the optical pulse waveforms to the R-peak, ensuring consistent alignment of the cardiac cycle across cortical locations. R-peaks were detected using a custom MATLAB script (2021B) that applied a dynamic voltage threshold and excluded peaks occurring outside the normal interbeat interval range. All detections were visually inspected, and artefacts (such as misclassified T-waves) were corrected or removed.

##### Structural Magnetic Resonance Imaging (MRI)

High-resolution T1-weighted anatomical images were acquired to co-register digitised source and detector coordinates to each participant’s T1-weighted anatomical image. MRI scans were obtained using a Siemens Prisma 3T scanner equipped with a 64-channel head and neck coil. T1-weighted anatomical images were acquired using a magnetisation-prepared rapid acquisition gradient echo (MPRAGE) sequence (acquisition time = 5:12 min; voxel size = 1×1×1 mm³; matrix size = 256×256×208).

Imaging parameters included a repetition time (TR) of 2300 ms, echo time (TE) of 2.91 ms, flip angle (FA) of 9°, and inversion time (TI) of 900 ms. Digitised coordinates for each participant were co-registered to their T1-weighted image using the Optimized Co-registration Package (OCP [33]) and optical data were subsequently combined with this co-registration solution during the optical analysis (Opt-3d processing [34]). In cases where individuals did not have either a digitisation or an MRI, co-registration was performed using a template.

##### Measuring PReFx over prefrontal cortex ROIs

Optical signals were normalised and band-pass filtered (0–10 Hz). Data were further analysed using Opt-3d software [34] to combine signals from channels with overlapping diffusion paths into weighted voxel-level waveforms. These signals were time-locked to the ECG R-wave. We report analyses using AC intensity values at 830 nm [18], resulting in 192 channels across the three montages.

Three large prefrontal cortex (PFC) regions of interest (ROIs) were defined using the Automated Anatomical Labelling Atlas 3 (AAL3 [35]): left PFC, middle PFC, and right PFC. The left and right PFC ROIs included the superior frontal gyrus, middle frontal gyrus, and dorsal segments of the inferior frontal gyrus (opercular and triangular parts). The middle PFC ROI included the bilateral superior frontal gyrus (medial part) and supplementary motor area (SMA) regions.

The waveform was normalised for pulse amplitude and time (systole-diastole interval). Following this, PReFx was calculated (in Opt-3d) as the area under the pulse waveform between systole and diastole, after subtracting a linear baseline. Individual trials (i.e., pulse cycles) for each ROI and each block were excluded if peaks could not be reliably detected, derived PReFx or amplitude values were outside plausible ranges, or optical interbeat interval estimates were < 40 bpm or > 120 bpm or inconsistent with ECG-derived values.

Individual trials (i.e., pulse cycles) for each ROI and rest block were excluded if (1) systolic or diastolic peaks could not be identified within the epoch, (2) PReFx was greater than 1.0 or less than -0.2, (3) peak amplitude was less than 0.001 dB or greater than 3 dB, (4) the interbeat interval estimated from the optical signal differed from the ECG-derived interbeat interval by more than 76.8 ms (3 sampling points), or (5) the interbeat interval was less than 500 ms (>120 bpm) or greater than 1500 ms (<40 bpm). For each ROI, at least 10% of voxels were required to have a minimum of 10 valid trials. For remaining trials, PReFx was calculated for each trial and then averaged across all the voxels in the ROI for each rest block. PReFx values from each rest block were averaged to obtain a single PReFx value for each ROI. To examine PReFx across the entire PFC, we also averaged the values from the three ROIs.

#### 2.2.3 Assessment of Cognitive Performance

Participants completed a battery of cognitive tasks to assess their performance in cognitive domains sensitive to ageing. Variables from these tasks were included in a confirmatory factor analysis (CFA) to derive three cognitive domain scores (Verbal Memory, Executive Function, and Processing Speed), as described below.

##### CANTAB Tasks

Three tasks from the Cambridge Neuropsychological Test Automated Battery were used (CANTAB) [36, 37]. The Verbal Recognition Memory (VRM) task assesses verbal memory and learning. The Multitasking Test (MTT) assesses set-shifting and interference control. The Reaction Time (RTI) task measures motor and mental response speed. As shown in Table 1, key outcome variables from each task were used to derive measures for each cognitive domain.

**Table 1.** Cognitive Domains, Tasks, and Outcome Measures.

| **Cognitive Domain** | **Task** | **Outcome Measure** |
| --- | --- | --- |
| *Verbal Memory* | CANTAB: | Immediate Free Recall |
|  | Verbal Recognition Memory | Immediate Recognition |
|  |  | Delayed Recognition |
| *Executive Function* | CANTAB: | MTT Multitasking Cost |
|  | Multitasking Test (MTT) | MTT Congruency Cost |
|  | Task-Switching Paradigm | TSWT Multitasking Cost |
|  | (TSWT) | TSWT Congruency Cost |
| *Processing Speed* | CANTAB: | Simple Reaction Time |
|  | Reaction Time task | Choice Reaction Time |

*Note.* TSWT = Task-Switching Paradigm; CANTAB =
Cambridge Neuropsychological Test Automated Battery;
TSWT Multitasking Cost is akin to what is typically
labelled general switch cost.

##### Cued-Trials Task-Switching Paradigm (TSWT)

The cued-trials task-switching paradigm (TSWT) assesses set-shifting and interference control (for details on task see [38, 39]. Briefly, participants completed simple classification tasks (i.e., letter: vowel or consonant; number: odd or even) in either single-task blocks (i.e., consecutive trials of same task) or mixed-task blocks (i.e., pseudorandom sequence alternating between two tasks). On each trial, a colour cue preceded the target that required a left- or right-hand response. The cue indicated whether to complete the letter or the number task (cue-target interval, CTI, 1000ms).

The target consisted of two characters, one of which was relevant to the cued task. The other character was either neutral (i.e., a non-alphanumeric character not mapped to any response) or an exemplar from the uncued task that is incongruently mapped to the response for the cued task. Reaction time and error rate are recorded. For each participant, trials with an incorrect response, and trials with reaction time (RT) below 200 ms or exceeding their condition mean by three standard deviations were excluded. TSWT Multitasking Cost is the difference between mixed-task and single-task blocks (i.e., general switch cost). TSWT Congruency Cost is the difference between incongruent and neutral trials (averaged over single-task and mixed-task blocks).

To account for potential speed-accuracy trade-off between reaction time and error rate on the MTT and TSWT tasks, we estimated Inverse Efficiency Scores (IES) for each trial type (i.e., mean RT on correct trials / (1 − error rate). Cost scores were derived from IES scores (with larger values reflecting greater cost/slower performance at derivation). Variable directionality was harmonised at the analysis stage so that higher values reflected better performance across cognitive indicators.

#### 2.2.4 Assessment of Covariates

##### Demographic variables

Demographic characteristics, including age (years), sex (male/female), marital status (single, de-facto, married, divorced, widowed), and employment status (including retirement status; yes/no), were obtained via self-report questionnaires. Total years of education was extracted from the Australian National University Alzheimer’s Disease Risk Index (ANU-ADRI) as the sum of primary, secondary, tertiary and ‘other’ education [40].

##### Total energy intake

Energy intake (kilojoules per day) was calculated from responses to the AES-FFQ using the AUSNUT 2011–2013 database to convert dietary data into mean daily nutrient and energy intake estimates [27, 30].

##### Moderate-to-vigorous physical activity (MVPA)

Daily MVPA time (minutes per day) was assessed using an Axivity AX3 triaxial accelerometer worn on the non-dominant wrist for seven consecutive days, with data recorded at 100 Hz. This device has good predictive validity for determining physical activity intensity and duration [41]. Raw data were downloaded using Open Movement GUI (OmGUI) software version 1.0.0.45 and further processed using a custom MATLAB R2018B interface (COBRA). Non-wear time was defined as ≥60 minutes of ≤25 g-min, verified against participant diaries and visual inspection of the accelerometry trace. From waking day activity traces, MVPA was classified as any waking acceleration exceeding >93 g-min [42]. Participants with at least three valid weekdays and one valid weekend day, and >16 hours of wear time per day were included in the analyses.

#### 2.2.5 Cardiometabolic Factors

##### Anthropometry

Weight and height were averaged over two measurements. Body Mass Index (BMI, weight (kg) / height (m)^2^) was calculated using the average of two height and weight measurements. Waist and hip circumference (cm) were measured to estimate waist-to-hip ratio (WHR).

##### Blood pressure and heart rate

Brachial blood pressure and heart rate measures were obtained by averaging over three measures from the left arm using an Omron automatic blood pressure monitor (model HEM-7000-CIL). Participants were seated for at least 5 minutes and had not consumed caffeine or tobacco in the previous hour.

##### Blood collection and analysis

Fasted blood samples were obtained via venipuncture. Samples were centrifuged at 4000 rpm for 10 minutes, aliquoted and stored at -80°C until further analysis. The KONELAB 20XTi auto-analyzer (ThermoFisher, Massachusetts, United States), with specific ThermoFisher reagents, performed analytical assessments of cholesterol (total and HDL), triglycerides, high-density lipoprotein (HDL) cholesterol, and glucose. Samples were thawed on ice, vortexed, and centrifuged at 10,000 rpm for two minutes to remove particulates. The instrument was calibrated using ThermoFisher calibrators. For each test run, 150 µL of the sample was transferred into a sample cup, ensuring that no bubbles were present. The autoanalyzer, with the necessary ThermoFisher reagents, analysed the samples based on light absorbency measurements. Low-density lipoprotein (LDL) cholesterol was calculated as LDL = total cholesterol – HDL – (trig/2.17).

##### Medication use

Participants were asked to bring in the packaging of their medications, and the formulation/type, dose, frequency, duration of use, were recorded by the study investigators.

### 2.3 Statistical Analysis

All analyses were conducted in R (version 4.4.2). Descriptive statistics are presented as means and standard deviations. Sex differences are tested using independent samples t-tests. Pearson correlation coefficients are used to assess relationships between variables of interest. Participants were grouped into quartiles based on the sample distribution of MedDiet score, and one-way ANOVAs were used to examine whether cardiometabolic and vascular outcomes differed across increasing levels of MedDiet adherence. Given the exploratory nature of this study, uncorrected p values are reported alongside Benjamini-Hochberg false discovery rate (FDR)-adjusted q values for the sex-difference, correlation, and quartile ANOVA analyses, with *q* < .05 used to indicate findings that survived correction.

Multiple linear regression models were estimated using complete cases for the variables included in each model (listwise deletion, n = 159), whereas the structural equation models (SEM) were estimated using FIML on the full Newcastle cohort (N = 198). The final sample can be seen in Figure 1. Analysis code is available at: https://github.com/f3lsimpson94/Mediterranean-Cerebral-Elasticity.

#### Primary Analysis: Association Between the MedDiet and Frontal Lobe Arterial Elasticity

The association between MedDiet and arterial elasticity (PReFx) was evaluated using multiple linear regression models. Model 1 included MedDiet as the predictor and PReFx as the outcome variable. Model 2 added demographic covariates (age, sex, education). Model 3 was further adjusted for lifestyle covariates (MVPA, total energy intake). Education was included as prior research has demonstrated controlling for education can attenuate the association between dietary patterns and cognition [43], while MVPA and energy intake were included to control for lifestyle factors influencing metabolic balance [44].

All continuous variables were standardised using z-score transformation (i.e., subtract sample mean and divide by standard deviation) to place them on a common scale and facilitate comparison of parameter estimates across variables. Multicollinearity among predictors was evaluated using variance inflation factors (VIFs) with the *car* package [45]. Standardised beta coefficients (β) and 95% confidence intervals (CIs) were computed using the package *QuantPsyc* [46]. Standardised 95% CIs for β were obtained using a delta-method calculation. For each linear regression model, we reported the multiple R² to indicate the proportion of variance in PReFx explained by the predictor(s). Regression assumptions (i.e., linearity, homoscedasticity, normality of residuals) were assessed via diagnostic plots.

Finally, we adopted exploratory analyses to examine whether the MedDiet-PReFx association was moderated by age, sex or MVPA, and the possibility of non-linear effects. Non-linearity was evaluated by including a quadratic (second-degree polynomial) term for MedDiet score in the regression model and by plotting and visually inspecting the association using a locally smoothed trend line. Influence diagnostics were also examined for the fully adjusted regression model (Model 3), and sensitivity refits excluding flagged cases are reported in Table S2 (Supplementary Material).

#### Secondary Analysis: Mediation Role of Cerebral Arterial Elasticity in the Association Between MedDiet and Cognitive Function

Cognitive performance was modelled with a confirmatory factor analysis (CFA) using the *lavaan* package [47] and involved a three-factor cognitive model adapted from Kałamała, Ware et al. [48].

Outlier screening was conducted for each of the nine cognitive indicators after *z*-standardisation. Univariate outliers (|*z*| > 3) were handled cell-wise by replacing extreme values with NA, while participants were retained. Multivariate outliers were then screened using a partial Mahalanobis procedure that tolerated partially missing data at the participant level and applied a chi-square cutoff based on the number of observed indicators available for each participant (See Supplementary Material for further information).

Before running the CFA, all continuous variables were standardised, and reaction time variables were reverse scored so that higher values reflected better performance. Missing data were handled using FIML. The proportion of missing data for individual variables ranged from 5% to 9%. All latent factors were specified as freely correlated. Equality constraints were applied to the Processing Speed indicators to improve model stability and interpretability [49], as the construct was measured by only two manifest highly correlated variables. Model fit was evaluated using the Comparative Fit Index (CFI), Tucker-Lewis Index (TLI), Parsimony Normed Fit Index (PNFI), Root Mean Square Error of Approximation (RMSEA), and Standardised Root Mean Square Residual (SRMR). Factor scores were extracted using lavPredict and used as outcome variables in the subsequent lavaan mediation models.

Separate mediation models were estimated in *lavaan* for Verbal Memory, Executive Function, and Processing Speed factors to test whether frontal lobe arterial elasticity (PReFx) mediated the association between the MedDiet and cognitive performance. Each model included path *a* (MedDiet -> PReFx), path *b* (PReFx -> cognitive outcome), and path *c*′ (direct effect of MedDiet on cognitive outcome, controlling for PReFx), and was adjusted for age, sex, education, MVPA, and total energy intake. The indirect effect was defined as *a × b*, and the total effect was estimated as *c*′ *+ (a × b)*. Models were estimated using FIML, and bootstrap resampling (1000 draws) was used to obtain confidence intervals for reported mediation parameters. Percentile bootstrap 95% confidence intervals were used to evaluate the indirect effects, with evidence of mediation inferred when the confidence interval did not include zero. Standardised estimates are reported.

## 3. Results

### Participant characteristics

The final sample consisted of 198 individuals with a mean age of 65.6 ± 3.2 years (see Figure 1). Of these, 58% were females, 76% were retired and 72% of participants were married. The average education level was 16.8 ± 3.2 years, reflecting a well-educated cohort. As detailed in Table 2, mean systolic blood pressure was just at the hypertensive range (140 mmHg), whereas diastolic blood pressure was in the healthy range as defined by the Heart Foundation of Australia [50]. Mean plasma total cholesterol level was at the upper limit of the healthy range [51], mean BMI was in the overweight category, consistent with Australian standards [52]. These cardiometabolic indicators align with mild-moderate cardiovascular risk profiles.

**Table 2.**
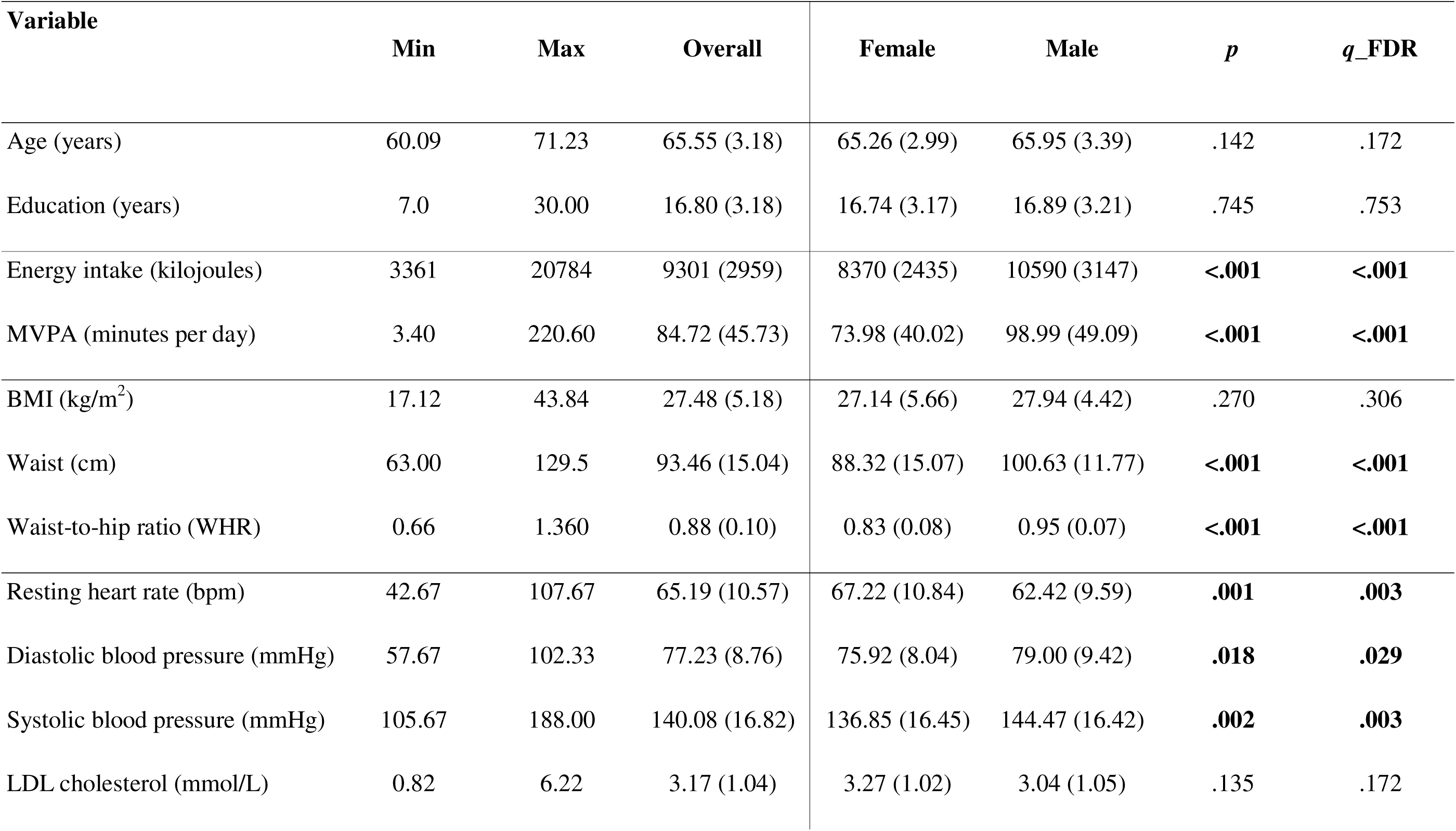
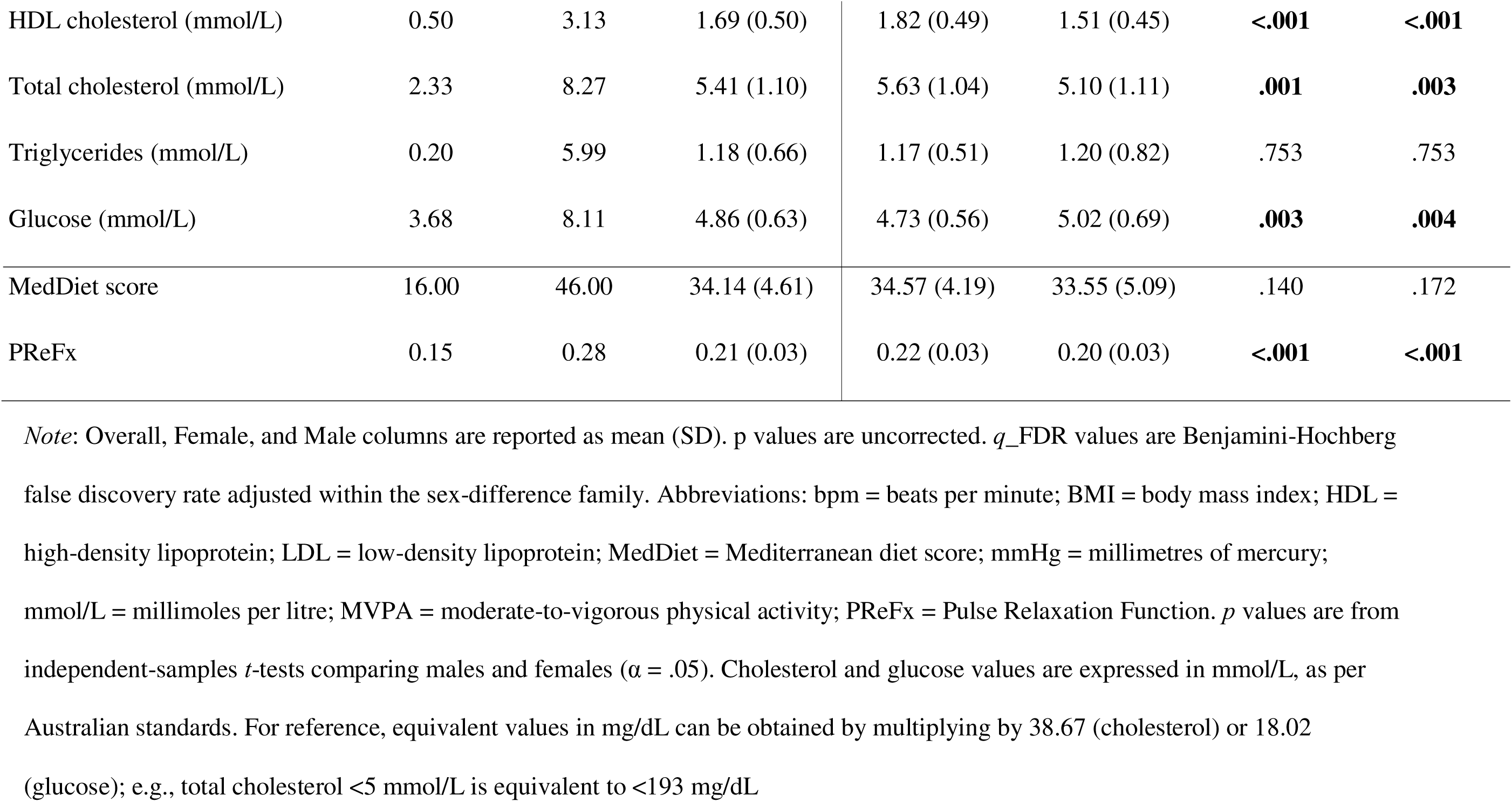
Demographic, cardiometabolic, and lifestyle characteristics by sex.

| Variable | Min | Max | Overall | Female | Male | *p* | *q_FDR* |
| --- | --- | --- | --- | --- | --- | --- | --- |
| Age (years) | 60.09 | 71.23 | 65.55 (3.18) | 65.26 (2.99) | 65.95 (3.39) | .142 | .172 |
| Education (years) | 7.0 | 30.00 | 16.80 (3.18) | 16.74 (3.17) | 16.89 (3.21) | .745 | .753 |
| Energy intake (kilojoules) | 3361 | 20784 | 9301 (2959) | 8370 (2435) | 10590 (3147) | **<.001** | **<.001** |
| MVPA (minutes per day) | 3.40 | 220.60 | 84.72 (45.73) | 73.98 (40.02) | 98.99 (49.09) | **<.001** | **<.001** |
| BMI (kg/m<sup>2</sup>) | 17.12 | 43.84 | 27.48 (5.18) | 27.14 (5.66) | 27.94 (4.42) | .270 | .306 |
| Waist (cm) | 63.00 | 129.5 | 93.46 (15.04) | 88.32 (15.07) | 100.63 (11.77) | **<.001** | **<.001** |
| Waist-to-hip ratio (WHR) | 0.66 | 1.360 | 0.88 (0.10) | 0.83 (0.08) | 0.95 (0.07) | **<.001** | **<.001** |
| Resting heart rate (bpm) | 42.67 | 107.67 | 65.19 (10.57) | 67.22 (10.84) | 62.42 (9.59) | **.001** | **.003** |
| Diastolic blood pressure (mmHg) | 57.67 | 102.33 | 77.23 (8.76) | 75.92 (8.04) | 79.00 (9.42) | **.018** | **.029** |
| Systolic blood pressure (mmHg) | 105.67 | 188.00 | 140.08 (16.82) | 136.85 (16.45) | 144.47 (16.42) | **.002** | **.003** |
| LDL cholesterol (mmol/L) | 0.82 | 6.22 | 3.17 (1.04) | 3.27 (1.02) | 3.04 (1.05) | .135 | .172 |
| HDL cholesterol (mmol/L) | 0.50 | 3.13 | 1.69 (0.50) | 1.82 (0.49) | 1.51 (0.45) | **<.001** | **<.001** |
| Total cholesterol (mmol/L) | 2.33 | 8.27 | 5.41 (1.10) | 5.63 (1.04) | 5.10 (1.11) | **.001** | **.003** |
| Triglycerides (mmol/L) | 0.20 | 5.99 | 1.18 (0.66) | 1.17 (0.51) | 1.20 (0.82) | .753 | .753 |
| Glucose (mmol/L) | 3.68 | 8.11 | 4.86 (0.63) | 4.73 (0.56) | 5.02 (0.69) | **.003** | **.004** |
| MedDiet score | 16.00 | 46.00 | 34.14 (4.61) | 34.57 (4.19) | 33.55 (5.09) | .140 | .172 |
| PReFx | 0.15 | 0.28 | 0.21 (0.03) | 0.22 (0.03) | 0.20 (0.03) | **<.001** | **<.001** |

*Note:* Overall, Female, and Male columns are reported as mean (SD). *p* values are uncorrected. *q*\_FDR values are Benjamini-Hochberg false discovery rate adjusted within the sex-difference family. Abbreviations: bpm = beats per minute; BMI = body mass index; HDL = high-density lipoprotein; LDL = low-density lipoprotein; MedDiet = Mediterranean diet score; mmHg = millimetres of mercury; mmol/L = millimoles per litre; MVPA = moderate-to-vigorous physical activity; PReFx = Pulse Relaxation Function. *p* values are from independent-samples *t*-tests comparing males and females ( $\alpha = .05$ ). Cholesterol and glucose values are expressed in mmol/L, as per Australian standards. For reference, equivalent values in mg/dL can be obtained by multiplying by 38.67 (cholesterol) or 18.02 (glucose); e.g., total cholesterol <5 mmol/L is equivalent to <193 mg/dL

The average MVPA was 85 min/day, indicating that, on average, participants maintained a highly active lifestyle. Approximately 26% of participants were taking antihypertensive medication, 18% were taking cholesterol-lowering medication and 6% were taking diabetes medication. Several sex differences remained after FDR correction.

Compared to females, males had higher energy intake, waist-to-hip ratio, systolic and diastolic blood pressure, and glucose levels, as well as lower HDL cholesterol, suggesting poorer cardiometabolic health (see Table 2). However, males also spent more time in MVPA, and had lower total cholesterol and resting heart rate than females.

### Mediterranean Diet Score

The mean MedDiet score was 34.14 (range: 16–46 out of 55), with a higher score indicating greater alignment with a MedDiet pattern. The mean MedDiet score did not differ across sexes (see Table 2). In bivariate analyses, higher MedDiet was associated with higher PReFx and lower BMI and waist circumference, and these correlations remained significant after FDR correction (Table 3). In contrast, none of the cardiometabolic or vascular variables differed significantly across MedDiet quartiles after FDR correction (Table S1, Supplementary Material).

**Table 3.** Bivariate Correlations with FDR Correction.

| Variable |  | Med | PReFx | Age | Edu | Energy | BMI | Waist | WHR | DP | SP | HR | Chol | HDL | LDL | Trigs | Gluc |
| --- | --- | --- | --- | --- | --- | --- | --- | --- | --- | --- | --- | --- | --- | --- | --- | --- | --- |
| PReFx | *r* | **.21** |  |  |  |  |  |  |  |  |  |  |  |  |  |  |  |
|  | *q* | **.020** |  |  |  |  |  |  |  |  |  |  |  |  |  |  |  |
| Age (years) | *r* | .06 | **-.30** |  |  |  |  |  |  |  |  |  |  |  |  |  |  |
|  | *q* | .568 | **<.001** |  |  |  |  |  |  |  |  |  |  |  |  |  |  |
| Education (years) | *r* | .08 | -.05 | .02 |  |  |  |  |  |  |  |  |  |  |  |  |  |
|  | *q* | .394 | .654 | .848 |  |  |  |  |  |  |  |  |  |  |  |  |  |
| Energy intake (kilojoules) | *r* | .10 | -.09 | -.01 | -.03 |  |  |  |  |  |  |  |  |  |  |  |  |
|  | *q* | .253 | .394 | .928 | .770 |  |  |  |  |  |  |  |  |  |  |  |  |
| BMI (kg/m<sup>2</sup>) | *r* | **-.18** | **-.23** | .02 | .04 | .06 |  |  |  |  |  |  |  |  |  |  |  |
|  | *q* | **.034** | **.012** | .884 | .726 | .593 |  |  |  |  |  |  |  |  |  |  |  |
| Waist (cm) | *r* | **-.20** | **-.33** | .05 | .05 | .17 | **.85** |  |  |  |  |  |  |  |  |  |  |
|  | *q* | **.017** | **<.001** | .625 | .654 | .053 | **<.001** |  |  |  |  |  |  |  |  |  |  |
| Waist-to-hip ratio (WHR) | *r* | -.15 | **-.41** | .04 | .07 | **.25** | **.43** | **.67** |  |  |  |  |  |  |  |  |  |
|  | *q* | .088 | **<.001** | .727 | .479 | **.002** | **<.001** | **<.001** |  |  |  |  |  |  |  |  |  |
| Diastolic blood pressure (BP) (mmHg) | *r* | -.04 | .02 | .04 | -.09 | .16 | **.22** | **.28** | **.24** |  |  |  |  |  |  |  |  |
|  | *q* | .725 | .902 | .737 | .320 | .072 | **.007** | **<.001** | **.005** |  |  |  |  |  |  |  |  |
| Systolic blood pressure (BP) (mmHg) | *r* | -.13 | **-.20** | **.29** | -.16 | .16 | .10 | **.19** | .17 | **.65** |  |  |  |  |  |  |  |
|  | *q* | .152 | **.040** | **<.001** | .079 | .082 | .277 | **.033** | .053 | **<.001** |  |  |  |  |  |  |  |
| Resting heart rate (HR) (bpm) | *r* | .00 | **-.28** | -.01 | .02 | -.12 | .17 | .09 | .12 | .09 | **-.16** |  |  |  |  |  |  |
|  | *q* | .965 | **.002** | .902 | .877 | .219 | .058 | .320 | .214 | .375 | **.069** |  |  |  |  |  |  |
| Total cholesterol (mmol/L) | *r* | .04 | .12 | -.11 | .09 | -.12 | -.16 | **-.22** | -.11 | -.01 | **-.08** | .02 |  |  |  |  |  |
|  | *q* | .726 | .223 | .223 | .338 | .194 | .086 | **.009** | .245 | .945 | **.391** | .896 |  |  |  |  |  |
| HDL cholesterol (mmol/L) | *r* | .13 | **.29** | -.01 | -.05 | -.08 | **-.44** | **-.56** | **-.45** | -.17 | **-.11** | -.04 | **.23** |  |  |  |  |
|  | *q* | .173 | **<.001** | .932 | .654 | .392 | **<.001** | **<.001** | **<.001** | .069 | **.232** | .737 | **.005** |  |  |  |  |
| LDL cholesterol (mmol/L) | *r* | -.01 | .05 | -.15 | .12 | -.10 | -.04 | -.07 | .02 | .03 | **-.08** | -.01 | **.92** | -.09 |  |  |  |
|  | *q* | .932 | .654 | .107 | .207 | .275 | .727 | .524 | .877 | .767 | **.442** | .928 | **<.001** | .371 |  |  |  |
| Triglycerides (mmol/L) | *r* | -.03 | **-.21** | .10 | .01 | .04 | **.29** | **.33** | **.27** | .14 | **.14** | .15 | .11 | **-.49** | .06 |  |  |
|  | *q* | .801 | **.027** | .286 | .943 | .708 | **<.001** | **<.001** | **<.001** | .137 | **.128** | .088 | .244 | **<.001** | .585 |  |  |
| Glucose (mmol/L) | *r* | -.13 | **-.30** | .13 | .02 | .03 | **.26** | **.35** | **.32** | .15 | **.22** | .04 | -.11 | **-.28** | -.07 | **.31** |  |
|  | *q* | .152 | **<.001** | .163 | .877 | .801 | **.001** | **<.001** | **<.001** | .113 | **.010** | .736 | .253 | **<.001** | .482 | **<.001** |  |
| MVPA (minutes per day) | *r* | .12 | **.19** | -.08 | .05 | **.19** | **-.36** | **-.28** | -.04 | -.11 | **.00** | **-.31** | .10 | **.29** | .03 | **-.23** | -.14 |
|  | *q* | .197 | **.048** | .442 | .654 | **.033** | **<.001** | **<.001** | .737 | .257 | **.995** | **<.001** | .320 | **<.001** | .805 | **.007** | .128 |

*Note.* Lower triangle shows Pearson correlation coefficients ( $r$ ) with Benjamini-Hochberg FDR-adjusted $q$ values directly below each $r$ value. **Bold values** indicate correlations surviving FDR correction ( $q < .05$ ). $N = 198$ . Correlations computed using pairwise complete observations.

**Table 4.** Linear regression models examining the association between MedDiet score and PReFx (n = 159)

|  | Variable | B | 95% CI | *p* |
| --- | --- | --- | --- | --- |
| Model 1 | MedDiet | 0.22 | 0.07, 0.38 | **.005** |
| Model 2 | MedDiet | 0.21 | 0.07, 0.35 | **.003** |
|  | Age | −0.28 | −0.42, −0.14 | **<.001** |
|  | Sex | −0.34 | −0.47, −0.20 | **<.001** |
|  | Education | −0.08 | −0.22, 0.06 | .244 |
| Model 3 | MedDiet | 0.16 | 0.03, 0.30 | **.019** |
|  | Age | −0.24 | −0.37, −0.11 | **.001** |
|  | Sex | −0.44 | −0.58, −0.29 | **<.001** |
|  | Education | −0.09 | −0.22, 0.04 | .161 |
|  | MVPA | 0.28 | 0.14, 0.42 | **<.001** |
|  | Energy intake | 0.01 | −0.13, 0.15 | .841 |

*Note.* MVPA = Moderate-to-vigorous physical activity; $\beta$ = standardised beta coefficients; 95% CI = standardised 95% confidence intervals. Significant *p*-values are bolded ( $p < .05$ ).

### Cognitive Performance

CFA was used to estimate three cognitive latent factors. Standardised loadings and model fit can be seen in Figure 2. Missingness was assessed using *TestMCARNormality* in R [53], and the non-parametric component of the test did not indicate a significant departure from data being missing completely at random (*p* = .383). FIML was therefore used to retain all available information when estimating the latent factors (see Supplementary Material Table S3 for summary of missing data per variable). The model demonstrated adequate fit (see Figure 2).

**Figure 2.**
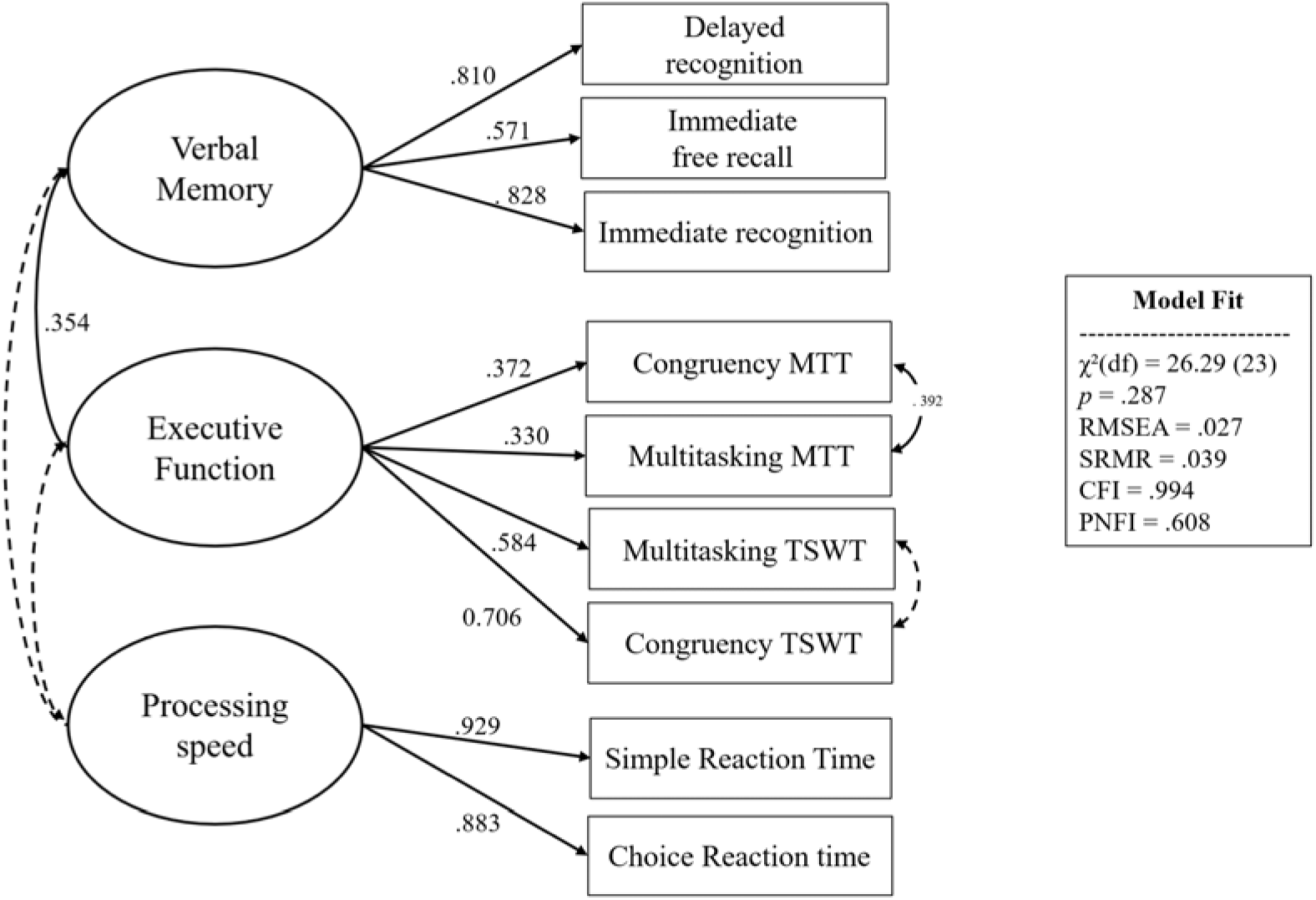
Confirmatory Factor Analysis (CFA) Model of Cognition *Note.* CFA model of cognitive latent variables estimated using FIML. After removal of three multivariate outliers, 195 participants were retained; five with no observed cognitive indicator data were omitted automatically by *lavaan*, resulting in 190 observations used for model estimation. Values shown on the single-headed arrows are standardised factor loadings. The Processing Speed factor loadings were constrained to be equal. Solid curved paths indicate statistically significant correlations or residual covariances, and dashed curved paths indicate non-significant correlations or residual covariances. All displayed factor loadings were statistically significant (*p* < .05).

### Cerebral Arterial Elasticity

Figure 3 shows that PReFx significantly decreased with increasing age, even within the restricted range of 60 to 70 years. PReFx was also significantly higher in females than males, consistent with previously reported sex differences in cerebral arterial elasticity [19] (Table 2). Higher PReFx was associated with lower BMI, waist circumference, WHR ratio, resting heart rate, and glucose levels, as well as higher HDL cholesterol.

**Figure 3.**
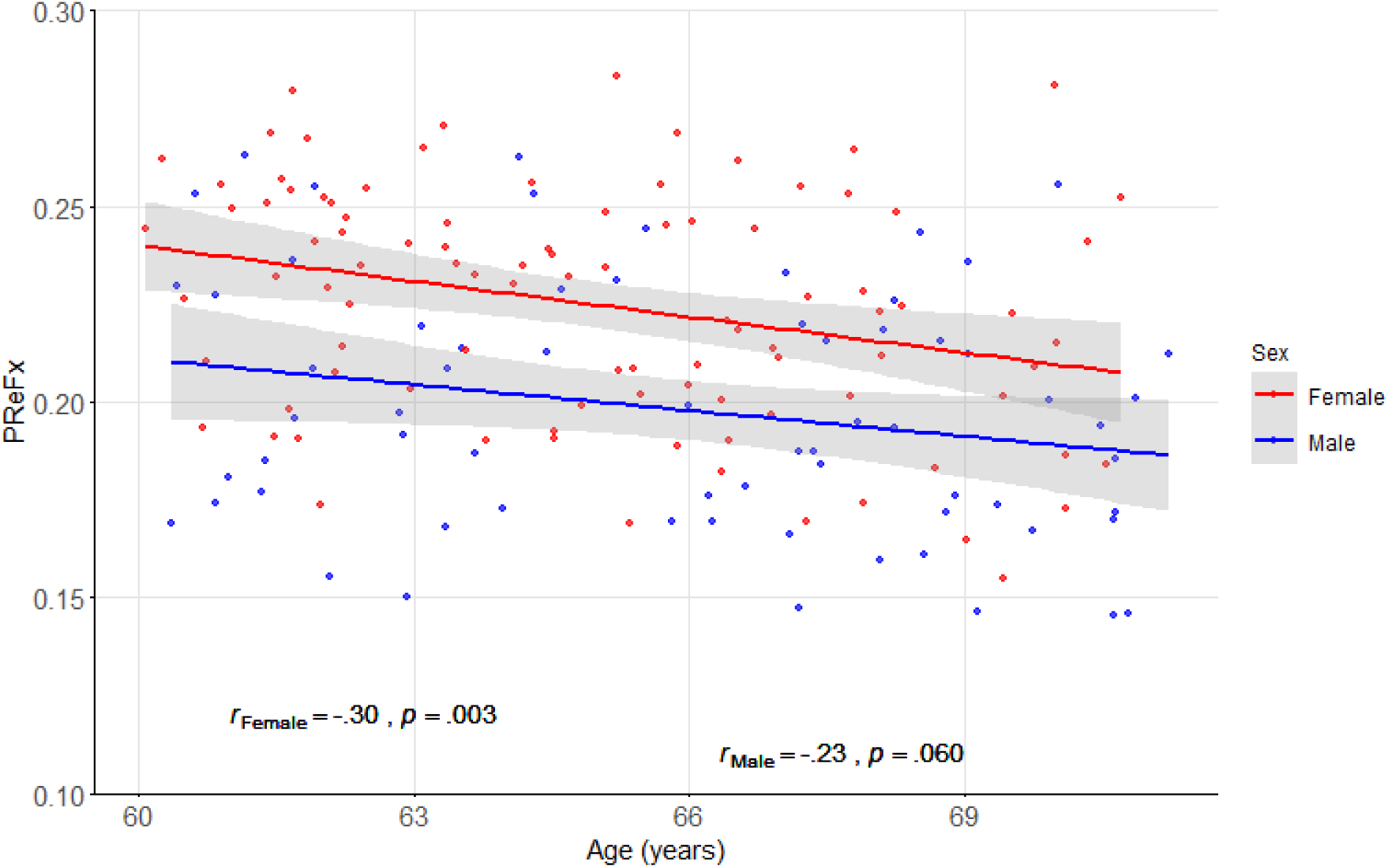
Correlation between Age and PReFx Stratified by Sex *Note*. The association between age and PReFx in females (red) and males (blue). Shaded regions represent 95% confidence intervals (95% CI). Females exhibited higher PReFx values than males across the age range, consistent with sex differences in cerebral arterial elasticity previously reported by Bowie and colleagues [19].

### Primary Analysis: MedDiet and Cerebral Arterial Elasticity

A higher MedDiet score was associated with higher PReFx in the unadjusted model, explaining 5% of the variance in PReFx, *F*(1, 157) = 8.16, *p* = .005. This association remained significant after adjustment for age, sex, and education, β = 0.21, 95% CI [0.07, 0.35], *p* = .003, with the model explaining 27% of the variance, *F*(4, 154) = 14.56, *p* < .001.

The association also remained significant in the fully adjusted model that further included MVPA and energy intake, β = 0.16, 95% CI [0.03, 0.30], *p* = .019, and the model explained 34% of the variance in PReFx, *F*(6, 152) = 13.29, *p* < .001 (Figure 4). In the fully adjusted model, higher MVPA was independently associated with higher PReFx (β = 0.28, *p* < .001), whereas energy intake was not associated with PReFx (β = 0.01, *p* = .841). Variance inflation factors (VIFs) were below five across all models, indicating no concerns with multicollinearity. Sensitivity refits excluding potentially influential cases yielded similar point estimates, supporting the robustness of the MedDiet-PReFx association (Table S2, Supplementary Material).

**Figure 4.**
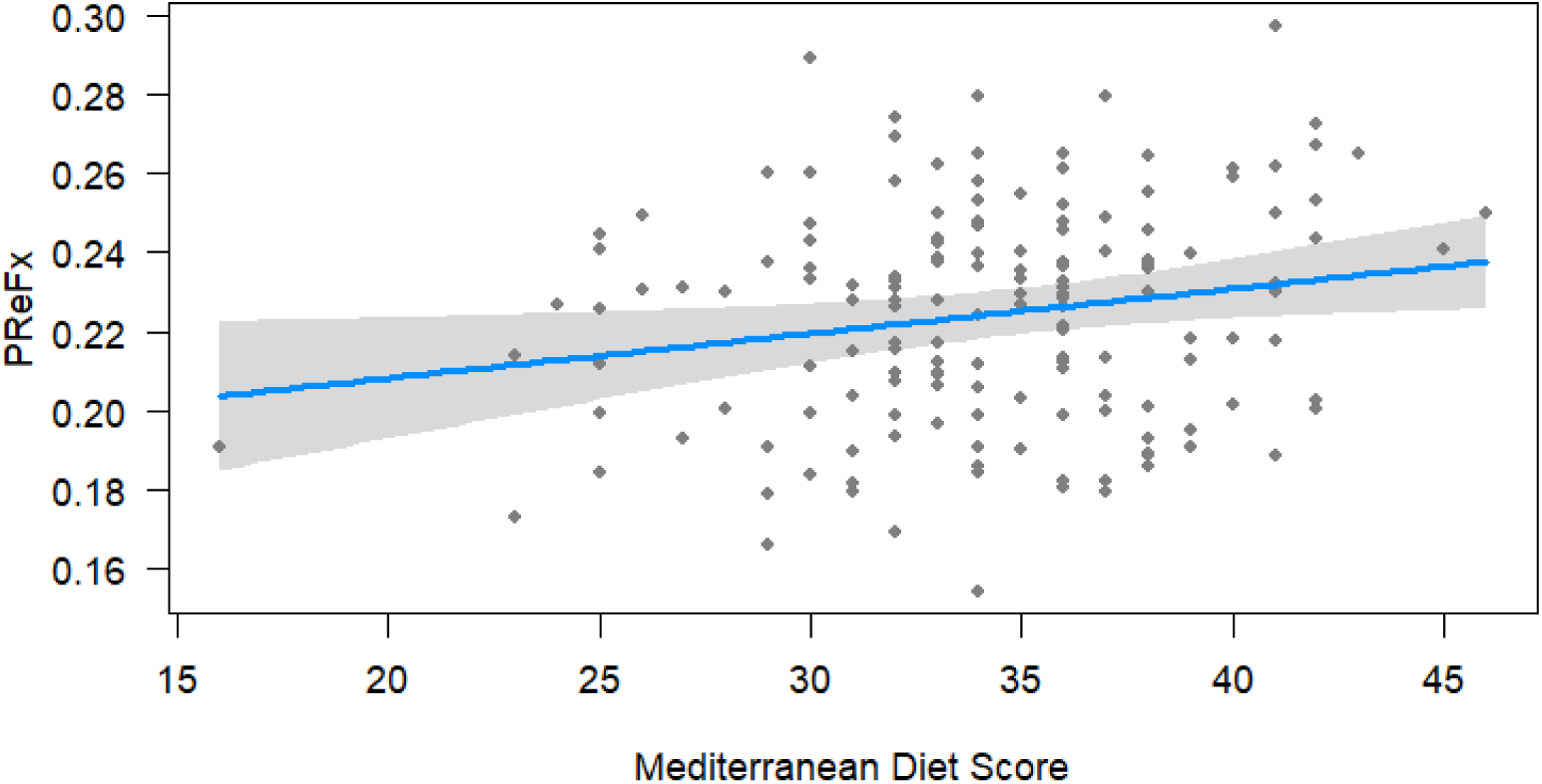
Association between the Mediterranean Diet and Cerebral Arterial Elasticity *Note*. Adjusted association between the Mediterranean diet and the Pulse Relaxation Function (PReFx), a measure of frontal lobe arterial elasticity. The solid blue line represents the predicted values from the fully adjusted regression model (Model 3), with shaded regions indicating the 95% confidence interval. Individual data points are shown in the background.

Exploratory post-hoc analyses examined whether the association between MedDiet and PReFx in Model 3 was moderated by age, sex or MVPA. None of these interaction terms were statistically significant. In addition, there was no evidence of non-linear effects as inclusion of a quadratic MedDiet term did not improve model fit, indicating that the association between MedDiet and PReFx was adequately captured by a linear model.

Figure 5 illustrates PFC arterial elasticity in participants in the lowest and highest MedDiet adherence quartiles, with warmer colours indicating lower arterial elasticity. The black line depicts the ROIs for left, middle and right PFC where PReFx was measured. For all three ROIs, participants with the highest MedDiet adherence showed greater arterial elasticity over the PFC, compared to those in the lowest adherence.

**Figure 5.**
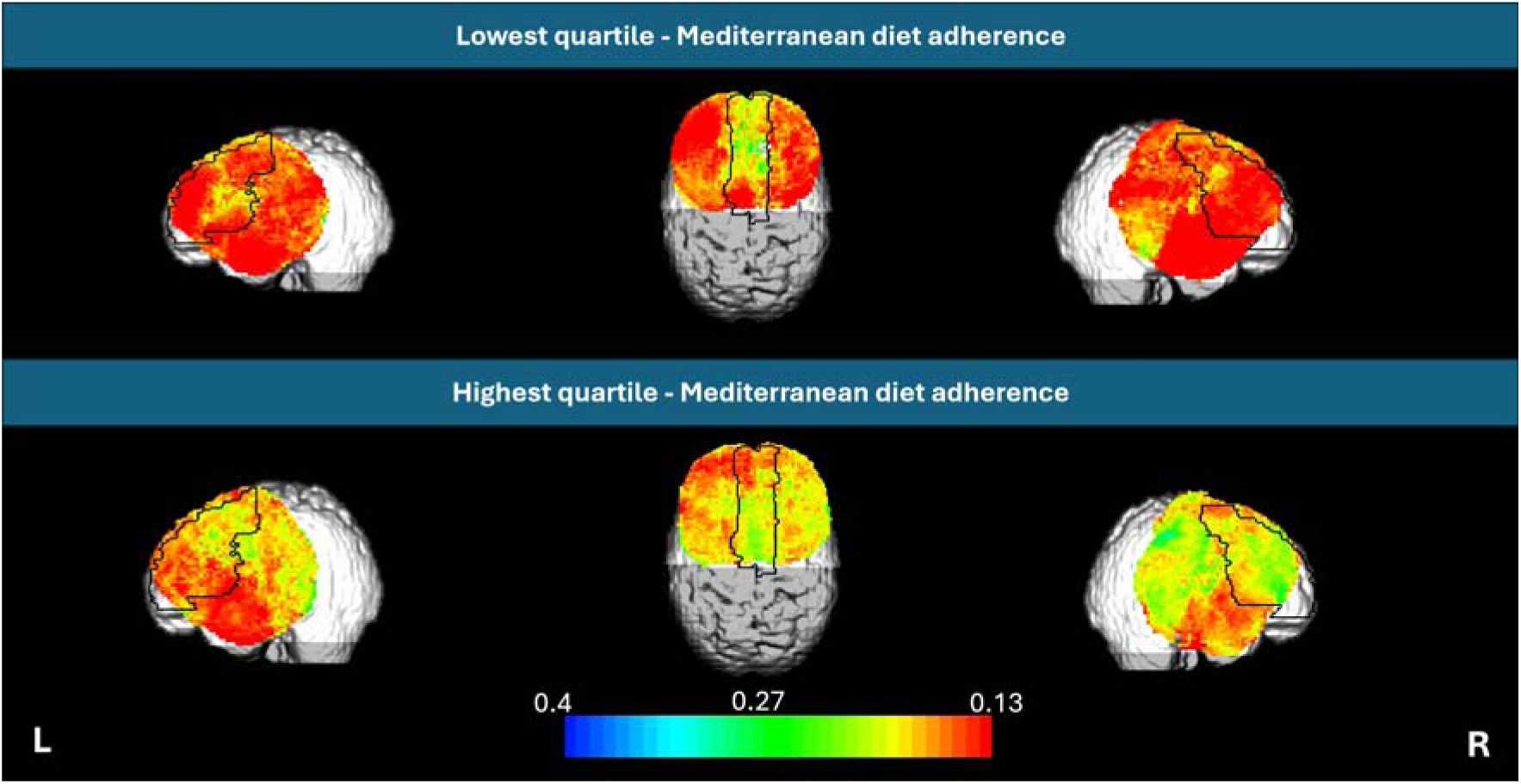
Cerebral Arterial Elasticity for participants in the lowest compared to the highest MedDiet adherence score quartile. *Note.* Pulse relaxation function (PReFx) maps created in Opt-3d [34] illustrate cerebral arterial elasticity in participants with the lowest (top row) and highest (bottom row) quartiles, according to MedDiet score. Quartiles were used for visualisation only (not inferential testing). The displayed maps represent group-averaged PReFx values within the lowest and highest MedDiet quartiles. Colour bar represents PReFx values, with cooler colours indicating greater elasticity (higher compliance) and warmer colours indicating reduced elasticity (lower compliance). L = Left; R = Right. Black outline depicts the PFC ROIs at which PReFx scores were measured.

### Secondary Analysis: Mediation of Cerebral Arterial Elasticity in the Association Between MedDiet and Cognitive Function

Separate mediation models were estimated for Verbal Memory, Executive Function, and Processing Speed to test whether prefrontal PReFx mediated associations between MedDiet score and cognitive performance. There was no statistically significant indirect effect for any cognitive domain, as the 95% confidence intervals for all indirect effects (a × b) included zero (Table 5). Direct effects (c′) and total effects were also non-significant across all three domains.

**Table 5.** Mediation analysis of the association between Mediterranean diet score and cognitive outcomes via PReFx (N = 198)

| Outcome | Effect Type | $\beta$ | 95% CI | *p* |
| --- | --- | --- | --- | --- |
| Verbal Memory | a path | 0.14 | 0.01, 0.26 | .030 |
|  | b path | -0.15 | -0.31, 0.03 | .117 |
|  | c' path (direct) | 0.13 | -0.02, 0.26 | .112 |
|  | a*b path (indirect) | -0.02 | -0.05, 0.00 | .221 |
|  | Total | 0.11 | -0.04, 0.23 | .165 |
| Executive Function | a path | 0.15 | 0.01, 0.27 | .025 |
|  | b path | 0.09 | -0.09, 0.20 | .374 |
|  | c' path (direct) | -0.03 | -0.14, 0.10 | .713 |
|  | a*b path (indirect) | 0.01 | -0.01, 0.04 | .439 |
|  | Total | -0.02 | -0.12, 0.11 | .831 |
| Processing Speed | a path | 0.14 | 0.01, 0.27 | .027 |
|  | b path | -0.05 | -0.21, 0.12 | .615 |
|  | c' path (direct) | -0.02 | -0.17, 0.12 | .800 |
|  | a*b path (indirect) | -0.01 | -0.04, 0.02 | .648 |
|  | Total | -0.03 | -0.17, 0.11 | .731 |

*Note.* $\beta$ values are standardised estimates. Confidence intervals are percentile bootstrap 95% confidence intervals (1,000 draws) from lavaan mediation models estimated with Full Information Maximum Likelihood (FIML). Models were fit on the full analytic cohort (N = 198); factor scores were observed for 190 participants, with the remaining 8 contributing through observed variables available under FIML
but not through the cognitive outcome itself. Mediation was inferred when the 95% confidence interval for the indirect effect ( $a \times b$ ) excluded zero.

## 4. Discussion

The current study demonstrated that greater alignment with the Mediterranean dietary pattern is associated with higher cerebral arterial elasticity. This relationship remained significant when controlling for demographic variables (age, sex, education), time spent in moderate-to-vigorous physical activity (MVPA), and energy intake. To our knowledge, this is the first study to demonstrate that a more healthful dietary pattern is associated with cerebral arterial elasticity over the PFC, an area known to be highly sensitive to ageing. Pulse-DOT offers a non-invasive, sensitive measure of cerebrovascular health that can detect subtle vascular changes before the emergence of brain structural damage [19], and has been associated with variability in cognitive abilities supported by these brain regions [23]. These findings support the notion that pulse-DOT measures may provide valuable insights into the impact of lifestyle factors on brain vascular health, well before the emergence of cognitive changes associated with increased dementia risk.

The present findings are consistent with a cerebrovascular pathway through which dietary patterns may contribute to neuroprotection. At the systemic level, arterial stiffening is associated with chronic inflammation, oxidative stress, and endothelial dysfunction, all of which can be modulated by dietary exposures (see reviews: [54, 55, 56]). The MedDiet pattern is characterised by higher intakes of monounsaturated and polyunsaturated fatty acids, fibre, and polyphenols, and has antioxidant and anti-inflammatory properties that may help preserve arterial function [56]. In contrast, dietary patterns characterised by refined carbohydrates and saturated fat contribute to an atherogenic profile, promoting arterial stiffening [58]. Given that cerebrovascular function relies on the integrity of small arteries and capillaries, dietary patterns that optimise systemic vascular health, such as the MedDiet, likely support the cerebrovasculature as well [59, 60].

Despite the significant association between the MedDiet and cerebral arterial elasticity, we found no significant relationship between diet and cognitive performance. This pattern is broadly consistent with literature showing that diet-cognition associations are often mixed and vary by cognitive domain, cohort characteristics, vascular risk, and analytic approach [2, 61, 62]. Moreover, several characteristics of the present cohort may have reduced sensitivity to detect subtle cross-sectional diet-cognition associations. The ACTIVate sample was cognitively intact and restricted to a relatively narrow age range (60 to 70 years). Participants had high educational attainment and were highly physically active, with an average MVPA of 85 minutes per day. Moreover, MedDiet scores were concentrated in the middle range of the scale (mean = 34.14, range: 16-46 out of 55), limiting the dietary contrast between participants and reducing statistical power to detect diet-cognition associations. It is possible that this relatively healthy, active sample was already near ceiling on the vascular pathways through which diet typically influences cognition, leaving less room for dietary variation to predict cognitive differences.

Diet-cognition associations also appear more robust in older ages or in cohorts with greater cardiometabolic burden. For example, the Maine-Syracuse Longitudinal Study reported MedDiet-cognition associations primarily in individuals aged over 70 years [63]. Similarly, studies recruiting participants with higher cardiovascular risk have more consistently reported diet-cognition associations [64, 65, 66]. Therefore, it may be that individuals with cardiometabolic dysfunction may derive greater cognitive benefit from dietary modification than metabolically healthy individuals. However, few studies have explored whether cardiometabolic risk moderates diet-cognition associations. In a cross-sectional study of older adults, Gauci and colleagues [67] found that dietary pattern adherence was not independently associated with cognitive performance. However, a prudent-style diet significantly moderated the association between arterial stiffness (carotid-femoral PWV) and verbal memory outcomes. This raises the possibility that diet may shape cognitive outcomes partly through its interaction with vascular risk, rather than acting as a strong independent predictor in relatively healthy samples.

Lastly, several limitations are worth noting. First, while the AES-FFQ is a well-validated tool, self-report dietary assessment is subject to recall biases, potentially leading to overestimation of socially desirable foods (e.g., vegetables, wholegrains) and underestimation of poorer-quality foods (e.g., processed foods) [68]. Second, the cross-sectional nature of this study means that temporal precedence cannot be established, and the observed diet-PReFx association may reflect shared underlying health behaviours rather than a directional effect of dietary pattern on arterial elasticity. Finally, while PReFx provides a non-invasive, highly reliable, regional index of cerebral arterial elasticity, replication across independent cohorts and integration with complementary vascular and neuroimaging markers will be important to establish convergent validity and clarify clinical relevance [18, 19, 20, 21, 22, 23].

Taken together, this study is the first to provide direct evidence that greater alignment with a Mediterranean dietary pattern is associated with greater prefrontal cerebral arterial elasticity in a sample of cognitively healthy older adults. These findings support a vascular pathway through which diet may influence brain ageing before the onset of overt cognitive decline, and highlight PReFx as a potentially useful outcome for future intervention studies. Both longitudinal and intervention studies are now needed to determine whether dietary modification can help maintain cerebral arterial elasticity and prevent or delay cognitive decline in older age.

## Acknowledgements

We thank all the people that supported the data administration and acquisition: Shania M Soman, Nathan Beu, Nathan Tran, Karen Wilson, Jodie Allen, Reuben Churcher, Jarvis Macleod, Sarah De Bock, Bridget Elliot, Christopher Brown, Helen Nicholas, Gemma Furuhashi Evans, Fayeem Bin Aziz, Mahmoud Abdolhoseini, Kate Dyer, Johanna Paddick, Ashlee Harvey, Leon Day, Elena Milochis, Alannah Graziano, Elizabeth Taddeo, Riley Jackson, Teigan Cotterill, Louise Massie, Mahmoud David Metherell. We thank the HMRI Research Volunteer Register for some participant recruitment.

## Author Contributions

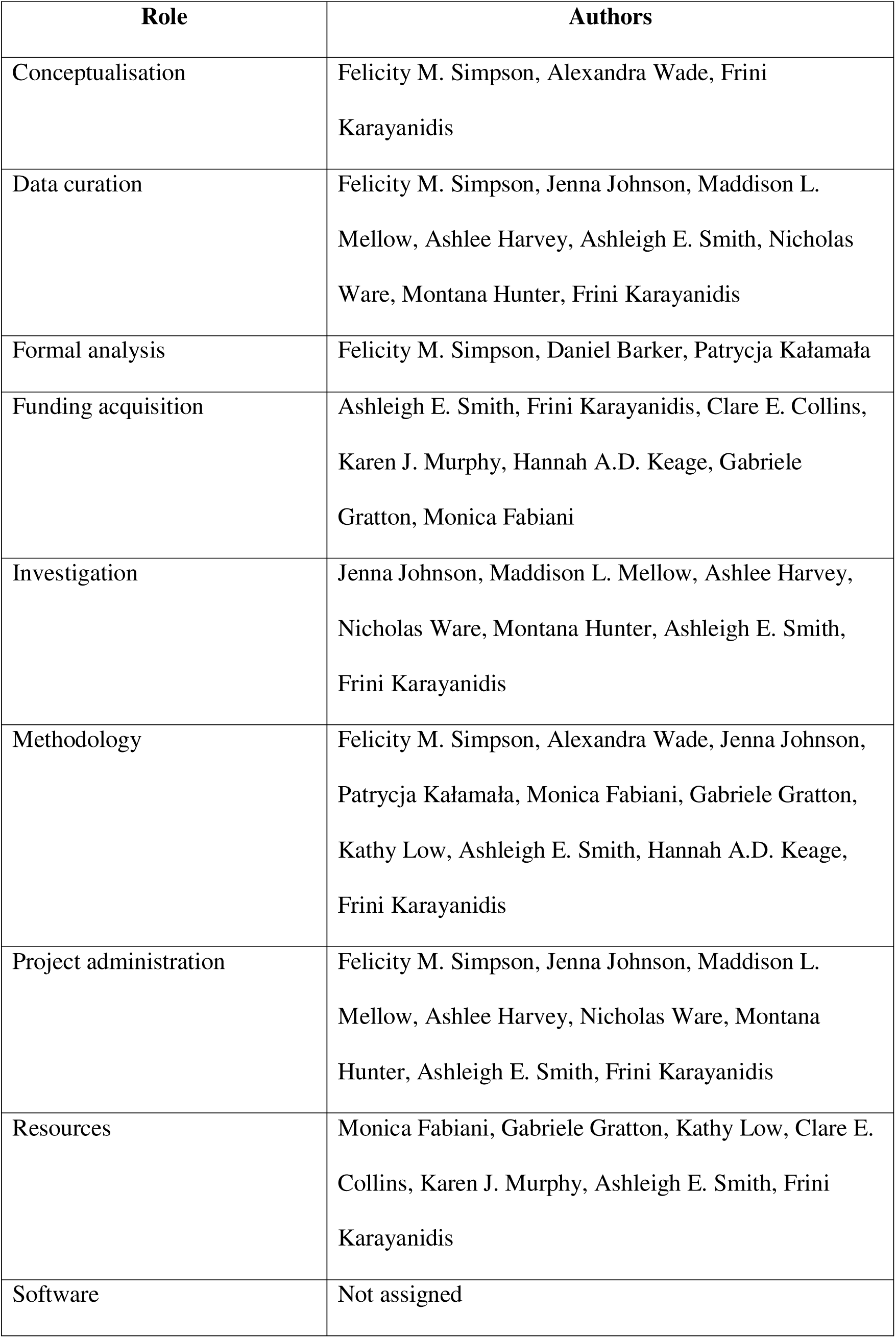
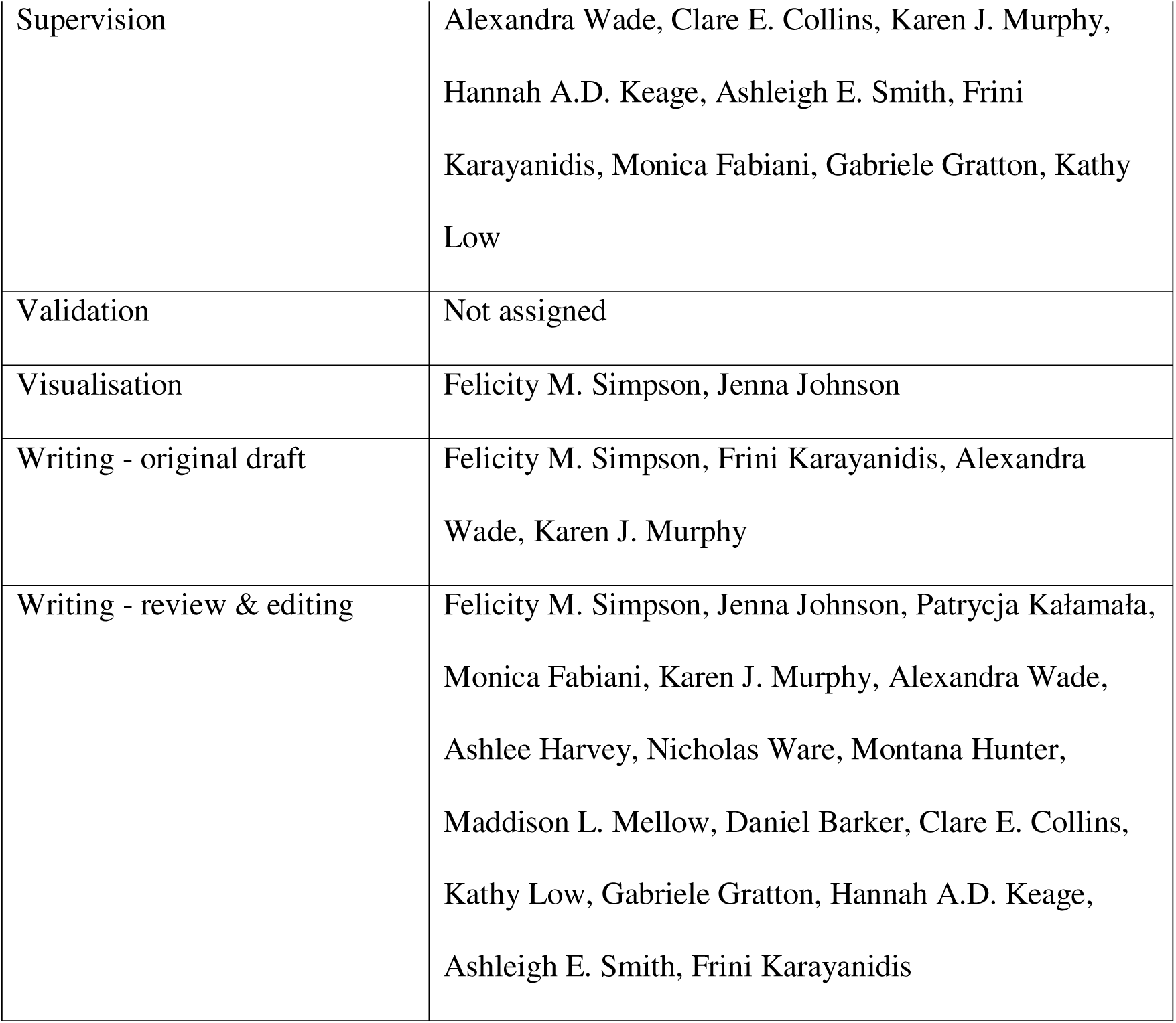

## Data Availability Statement

The data that support the findings of this study are not publicly available because they are derived from human participant data from the ACTIVate study and are subject to ethics and privacy restrictions. De-identified data may be made available by the corresponding authors on reasonable request, subject to approval by the relevant Human Research Ethics Committees and any institutional data access requirements. Analysis code used in this study is publicly available in the GitHub repository cited in the manuscript.

