## Supplementary Material for "The Mediterranean Diet is Associated with Higher Arterial Elasticity over Prefrontal Cortex in Older Adults"

**Table S1.** MedDiet quartile ANOVAs with FDR correction

| **Variable** | **Q1 (16-32)** n = 50 | **Q2 (>32-34)**  n = 56 | **Q3 (>34-37)**  n = 45 | **Q4 (>37-47)**  n = 47 | ***P*** | ***q*_FDR** |
| --- | --- | --- | --- | --- | --- | --- |
| Age (years) | 65.59 (3.00) | 65.35 (3.14) | 65.09 (3.33) | 66.19 (3.24) | .379 | .608 |
| BMI (kg/m2) | 28.43 (4.84) | 28.44 (5.49) | 26.41 (5.35) | 26.41 (4.69) | .058 | .370 |
| Education (total years) | 16.69 (3.59) | 16.56 (2.89) | 16.91 (3.15) | 17.11 (3.16) | .840 | .979 |
| Resting heart rate (bpm) | 64.59 (10.46) | 65.22 (10.49) | 65.11 (9.52) | 65.83 (11.97) | .957 | .979 |
| Energy intake (kilojoules) | 8804 (2976) | 9610 (3362) | 8753 (2792) | 9986 (2439) | .107 | .374 |
| MVPA (minutes) | 72.71 (39.55) | 88.90 (48.41) | 91.11 (43.40) | 86.66 (49.79) | .191 | .420 |
| Waist-to-hip ratio | 0.90 (0.10) | 0.88 (0.09) | 0.84 (0.08) | 0.89 (0.11) | **.032** | .370 |
| Diastolic blood pressure (mmHg) | 77.02 (7.91) | 78.45 (9.23) | 76.67 (8.70) | 76.50 (9.19) | .663 | .928 |
| Systolic blood pressure (mmHg) | 141.54 (16.59) | 143.72 (15.89) | 138.94 (17.51) | 135.36 (16.78) | .079 | .370 |
| LDL cholesterol (mmol/L) | 3.14 (1.010) | 3.22 (1.26) | 3.15 (1.00) | 3.16 (0.83) | .979 | .979 |
| HDL cholesterol (mmol/L) | 1.66 (0.55) | 1.58 (0.46) | 1.76 (0.45) | 1.78 (0.51) | .175 | .420 |
| Total cholesterol (mmol/L) | 5.37 (1.04) | 5.30 (1.34) | 5.53 (1.03) | 5.45 (0.93) | .778 | .979 |
| Triglycerides (mmol/L) | 1.23 (0.69) | 1.08 (0.44) | 1.33 (0.90) | 1.10 (0.51) | .210 | .420 |
| Glucose (mmol/L) | 4.93 (0.66) | 4.92 (0.59) | 4.75 (0.67) | 4.79 (0.62) | .391 | .608 |

*Note.* Values are mean (SD). p values are uncorrected omnibus one-way ANOVA p values. *q*_FDR values are Benjamini-Hochberg false discovery rate adjusted within the quartile ANOVA family. No quartile omnibus ANOVA remained significant after FDR correction.

**Outlier Screening**

Cognitive indicators were screened for outliers in two stages after z-standardisation. First, univariate outliers were identified cell-wise as values with absolute z-scores exceeding 3. These cells were replaced with NA while the participant was retained, resulting in 16 cells replaced across 14 participants. Second, multivariate outliers were identified using a partial Mahalanobis procedure that accommodated partially missing rows by computing Mahalanobis distances using only the observed indicators for each participant, provided at least three indicators were observed. A row-specific chi-square cutoff was applied at *p* = .003 (99.7th percentile), with degrees of freedom equal to the number of observed indicators for that participant. Three participants were identified as multivariate outliers and removed, leaving a retained sample of 195 participants for the CFA.

**Influential Cases**

We conducted regression influence diagnostics for the multiple linear regression model predicting arterial elasticity (PReFx) from Mediterranean diet and covariates (Model 3). Cook’s Distance, leverage, and DFBETAS were calculated for all observations. Cases exceeding conventional thresholds (Cook’s Distance > 4/n, leverage > 2×mean leverage, DFBETAS > 2/√n) were flagged as potentially influential. In Model 3 (*n* = 159), 8 cases exceeded the Cook’s distance threshold (4/*n*), 10 exceeded the leverage threshold (2×mean(hat)), and 35 exceeded the DFBETAS threshold (2/√*n*) when evaluated across all coefficients; 10 exceeded the DFBETA threshold for the MedDiet term specifically. The union of these criteria identified 42 unique cases. Sensitivity refits excluding flagged cases are shown in Table S2; point estimates were similar, so all observations were retained for the primary analyses.

**Table S2.** *Sensitivity Analyses for the MedDiet: PReFx Association (Model 3)*

| Model | n | β | 95% CI | *p* |
| --- | --- | --- | --- | --- |
| Full | 159 | 0.16 | [0.03, 0.29] | .019 |
| CookOnly | 151 | 0.14 | [0.01, 0.27] | .040 |
| LevOnly | 149 | 0.14 | [0.00, 0.28] | .057 |
| DFBETA_Med | 149 | 0.11 | [−0.03, 0.24] | .114 |
| Sans_All | 117 | 0.13 | [0.00, 0.27] | .057 |

*Note.* Values are standardised coefficients (Std β with 95% CIs); Rows indicate refits excluding: CookOnly = cases with Cook’s Distance > 4/*n*; LevOnly = leverage > 2×mean leverage; DFBETA_Med = |DFBETA| for the MedDiet term > 2/√*n*; Sans_All = union of all flags. CI = confidence interval.

**Table S3** *Missing Data Summary for Secondary Analysis Cognitive Indicators*

| **Variable** | **Missing (n)** | **Missing (%)** | **Valid (n)** |
| --- | --- | --- | --- |
| Delayed Recognition | 15 | 7.58% | 183 |
| Immediate Free Recall | 10 | 5.05% | 188 |
| Immediate Recognition | 10 | 5.05% | 188 |
| Simple-Choice Reaction Time | 12 | 6.06% | 186 |
| Five-Choice Reaction Time | 12 | 6.06% | 186 |
| MTT Congruency Cost | 12 | 6.06% | 186 |
| MTT Multitasking Cost | 12 | 6.06% | 186 |
| TSWT Congruency Cost | 18 | 9.09% | 180 |
| TSWT Multitasking Cost (General Switch Cost) | 18 | 9.09% | 180 |

*Note.* Table S3 presents the extent of missing data across cognitive outcome variables used in the Confirmatory Factor Analysis (CFA) model. Indicators span three cognitive domains: Verbal Memory (Delayed Recognition, Immediate Free Recall, Immediate Recognition), Executive Function (MTT Congruency Cost, MTT Multitasking Cost, TSWT Congruency Cost, TSWT Multitasking Cost), and Processing Speed (Simple-Choice Reaction Time, Five-Choice Reaction Time). Missingness per variable ranged from 5.05% to 9.09%. Little's MCAR test did not reject the assumption that data were Missing Completely at Random, *p* = .383, supporting the use of Full Information Maximum Likelihood (FIML) estimation to handle missing data in the CFA. Prior to model estimation, all cognitive indicators were z-standardised, and reaction time and cost variables were reverse-scored so that higher scores reflected better cognitive performance.
